# Longitudinal Changes in Functional Connectivity in Antipsychotic-treated and Antipsychotic-naive Patients with First Episode Psychosis

**DOI:** 10.1101/2021.04.13.21255375

**Authors:** Sidhant Chopra, Shona M. Francey, Brian O’Donoghue, Kristina Sabaroedin, Aurina Arnatkeviciute, Vanessa Cropley, Barnaby Nelson, Jessica Graham, Lara Baldwin, Steven Tahtalian, Hok Pan Yuen, Kelly Allott, Mario Alvarez-Jimenez, Susy Harrigan, Christos Pantelis, Stephen J Wood, Patrick McGorry, Alex Fornito

## Abstract

**Background:** Altered functional connectivity (FC) is a common finding in resting-state functional Magnetic Resonance Imaging (rs-fMRI) studies of people with psychosis, yet how FC disturbances evolve in the early stages of illness, and how antipsychotics may influence the temporal evolution of these disturbances, remains unclear. Here, we scanned first episode psychosis (FEP) patients who were and were not exposed to antipsychotic medication during the first six months of illness at baseline, three months, and 12 months, to characterize how FC changes over time and in relation to medication use.

**Methods:** Sixty-two antipsychotic-naïve patients with FEP received either an atypical antipsychotic or a placebo pill over a treatment period of 6 months. Both FEP groups received intensive psychosocial therapy. A healthy control group (n=27) was also recruited. A total of 202 rs-fMRI scans were obtained across three timepoints: baseline, 3-months and 12-months. Our primary aim was to differentiate patterns of FC in antipsychotic-treated and antipsychotic-naive patients within the first 3 months of treatment, and to examine associations with clinical and functional outcomes. A secondary aim was to investigate long-term effects at the 12-month timepoint.

**Results:** At baseline, FEP patients showed widespread functional dysconnectivity in comparison to controls, with reductions predominantly affecting interactions between the default mode network (DMN), limbic systems, and the rest of the brain. From baseline to 3 months, patients receiving placebo showed increased FC principally within the same systems, and some of these changes correlated with improved clinical outcomes. Antipsychotic exposure was associated with increased FC primarily between the thalamus and the rest of the brain. At the 12-month follow-up, antipsychotic treatment was associated with a prolonged increase of FC primarily in the DMN and limbic systems.

**Conclusions and Relevance:** Antipsychotic-naïve FEP patients show widespread functional dysconnectivity at baseline, followed by an early normalization of DMN and paralimbic dysfunction in patients receiving a psychosocial intervention only. Antipsychotic exposure is associated with distinct FC changes, principally concentrated on thalamo-cortical and limbic networks.

## Introduction

The symptoms of psychosis are widely thought to emerge from disrupted communication between large-scale functional brain networks^1,2^. Resting-state functional magnetic resonance imaging (rs-fMRI) has been widely used to map altered functional connectivity (FC) in patients across different illness stages^3–5^, and within specific cortico-subcortical^1,3^ and cortico-cortical networks ^6,7^. Numerous causes for altered FC in psychosis patients have been proposed, including abnormal neurodevelopment^8^, dopamine dysfunction^9^, traumatic early-life experiences^10^, and the neuromodulatory effect of antipsychotic medication^11^. In particular, widespread and early treatment of first episode psychosis (FEP) patients with antipsychotics has made it difficult to disentangle the effects of medication from other factors such as illness progression or the effects of non-pharmacological treatments on FC.

Cross-sectional studies have reported widespread hypoconnectivity in antipsychotic-naïve patients compared to healthy controls, with decreased connectivity between subcortical, frontal and temporal regions^12,13,14^. Hyperconnectivity has also been reported, with several studies showing increased connectivity between the thalamic and primary sensory cortices^16^ and within the default mode network^15,16,17^. These results suggest that not all FC abnormalities can be explained by medication. Nonetheless, medication does appear to affect FC, with longitudinal showing evidence of FC normalisation after a period of antipsychotic exposure^11^, particularly within fronto-thalamo-striatal circuits^12,18,19^, cortico-limbic ^18,20,21^, and cortico-cortical systems^5,22^, and which are sometimes correlated with symptom improvement^5,18^.

Unfortunately, no prior study has compared longitudinal FC changes in medicated and unmedicated patients, making it impossible to disentangle FC changes attributable to antipsychotic medication from those attributable to other factors such as natural course of the illness or adjunct interventions. This can only be done through a randomised, placebo-controlled study of antipsychotic-naïve patients. We recently used such a design^23^ to show that medicated and unmedicated patients show differential trajectories of grey matter volume, and that that antipsychotics may normalise basal ganglia volume in early illness stages.

Here, we report an analysis of FC in this cohort. FEP patients were scanned at baseline while antipsychotic-naïve and then again at 3 and 12 months after being randomized to receive treatment with either antipsychotic medication plus intensive psychosocial treatment (MIPT) or placebo plus intensive psychosocial treatment (PIPT). Our primary aim was to compare the evolution of FC changes over time in antipsychotic-treated patients compared to antipsychotic-naïve patients during the initial stages of psychotic illness. We also examined whether any observed FC changes were associated with changes in psychiatric symptoms and functional outcomes. In a secondary analysis, we investigated longer-term FC changes at the 12-month follow-up, after a period of time in which both groups had been exposed to antipsychotics. Critically we used a connectome-wide analysis to comprehensively map FC changes in antipsychotic-treated and antipsychotic-naïve patients at each time point.

## Method

### Study Design

The study took place at the Early Psychosis Prevention and Intervention Centre, which is part of Orygen Youth Health, Melbourne, Australia. The study was registered with the Australian New Zealand Clinical Trials Registry in November 2007 (ACTRN12607000608460) and received ethics approval from the Melbourne Health Human Research and Ethics committee. Patients were randomized to one of two groups: one given antipsychotic medication plus intensive psychosocial therapy (MIPT) and the other given a placebo plus intensive psychosocial therapy (PIPT). A third healthy control group who received no intervention was also recruited. For both patient groups, the treatment period spanned six months. MRI and clinical assessments were conducted at baseline, three months, and a final follow-up at 12 months. The randomization phase of the study terminated at 6 months, so patients in either the MIPT or PIPT group could have received antipsychotic medication and ongoing psychosocial interventions in between 6 and 12 months into the study. Further research and safety protocols can be found in the Supplement and elsewhere^24^.

### Participants

Patients were aged 15-25 years and were experiencing a first episode of psychosis, defined as fulfilling Structured Clinical Interview for DSM-IV (SCID) criteria for a psychotic disorder, including schizophrenia, schizophreniform disorder, delusional disorder, brief psychotic disorder, major depressive disorder with psychotic symptoms, substance-induced psychotic disorder or psychosis not otherwise specified. Additional inclusion criteria to minimise risk were: ability to provide informed consent; comprehension of English language; no contraindication to MRI scanning; duration of untreated psychosis (DUP) of less than 6 months; living in stable accommodation; low risk to self or others; none or minimal previous exposure to antipsychotic medication (< 7 days of use or lifetime 1750mg chlorpromazine equivalent exposure; further details provided in Supplementary table 1). A detailed participant-flow diagram can be found in Supplement.

### MRI acquisition and processing

Whole-brain T2*-weighed echo-planar images and anatomical T1-weighted (T1w) scans were acquired for each participant using a 3T Siemens Trio Tim scanner, equipped with a 32-channel head coil, located at the Royal Children’s Hospital in Melbourne, Australia. A total of 202 rs-fMRI datasets were acquired in this study, with rigours quality control procedures and standard image processing procedures used. A total of 193 scans survived our quality control procedure and to generate whole-brain FC matrices, we parcellated each individual’s normalised scans into 300 cortical^29^ and 32 subcortical regions^30^. Further details on acquisition, quality control and processing can be found in the Supplement.

### Statistical analyses

Non-parametric mixed-effects marginal models were used to analyse brain-wide FC changes across the three groups (MIPT, PIPT and controls) and three timepoints (baseline, 3-months and 12-months). The Network Based Statistic^123^ (NBS) was used to perform family-wise error-corrected (FWE) inference at the level of connected-components of edges, with the primary component-forming threshold, *τ*, set to *p* < .05. Further statistical details and results for *p* < .01 and *p* < .001 reported in the Supplement.

We evaluated NBS results using a Bonferroni-corrected threshold of *p*_*FWE*_ < .016, adjusted for three primary contrasts: (1) a contrast assessing baseline differences between healthy controls and patients; (2) a contrast isolating differential FC changes over time in the antipsychotic-naïve patient (PIPT) group, compared to the healthy control group; and (3) a contrast isolating the specific effects of antipsychotic treatment by examining differential FC changes over time in the antipsychotic-treated (MIPT) group, compared to both the PIPT and healthy control group.

For the secondary analysis of longer-term antipsychotic-naive and antipsychotic-related effects after the trial treatment period, we largely repeated the same procedure above, but focusing instead on differences between the baseline and 12-month timepoints. We used non-parametric canonical correlation analysis (CCA) to investigate the relationships between FC changes (ΔFC) within any identified NBS subnetworks and changes in the two pre-registered^24^ primary trial outcome measures––SOFAS and BPRS total scores^34^––across all patients. We summarized high-dimensional edgewise ΔFC values using principal component analysis (PCA). The threshold for statistical significance was set to *p*_*fwe*_ < .0125, which is Bonferroni-corrected for four CCA analyses. Further details are in the Supplement.

## Results

### Demographics and clinical characteristics

We have previously reported the demographics and clinical characteristics of this cohort^23^. Briefly, we detected no significant differences between the patient and control samples in sex or handedness, but the patients were, on average, 1.9 years younger and had 2 years less education (see Supplement for further details).

### Antipsychotic-naïve effects

#### Baseline

We identified a single NBS component showing widespread FC differences in patients compared to controls, comprising 4087 edges linking all 316 regions (*p*_*FWE*_ = .0152). Within this network, 54% (2195) of edges showed lower FC and 46% (1892) of edges showed higher FC in patients (Fig 1a-b).

**Figure 1.**
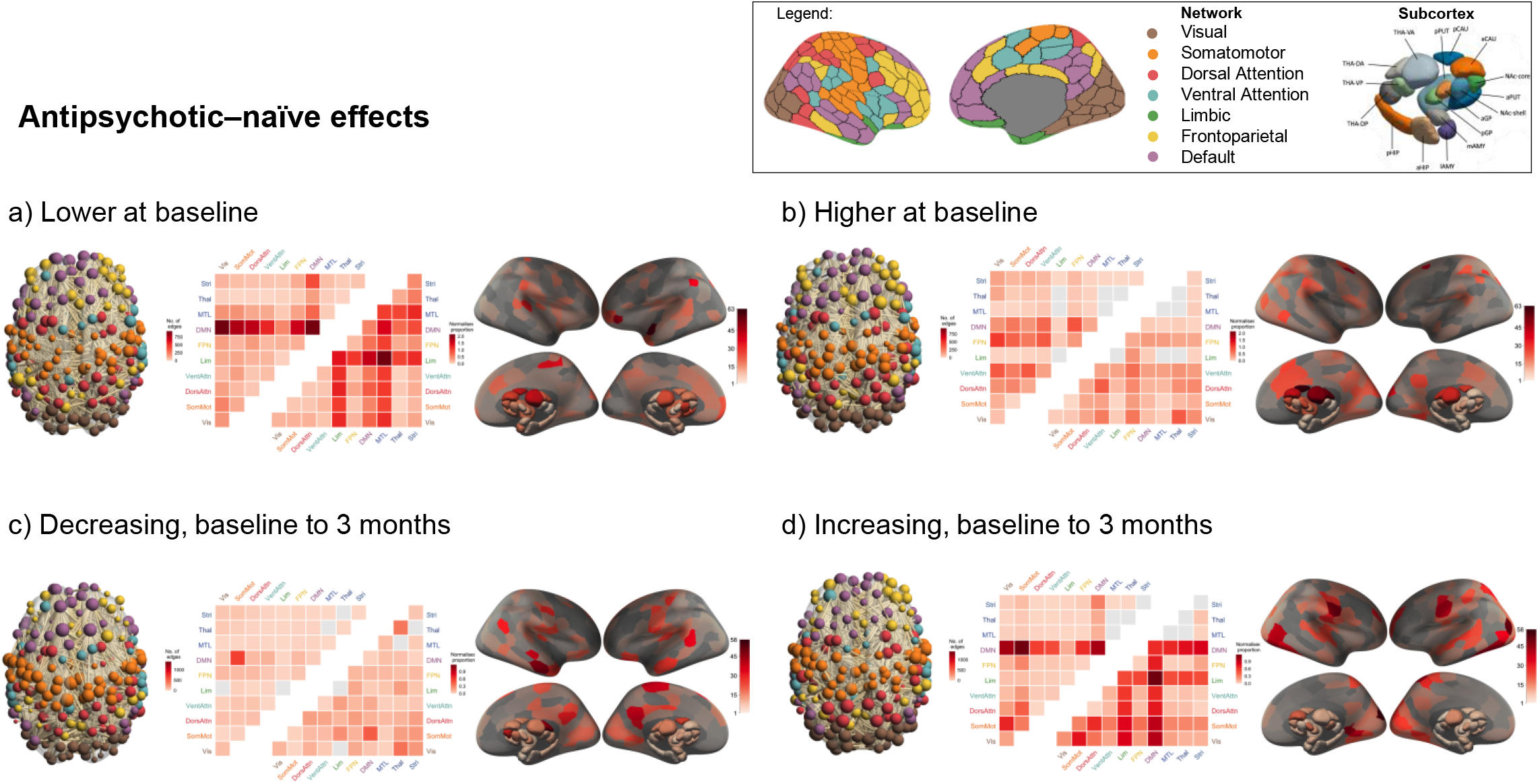
Baseline and short-term longitudinal functional connectivity changes in antipsychotic-naïve patients. Legend shows the Schaefer network parcellation^29^ and the Tian subcortex parcellation^30^ (Scale II). Each of the four panels contains three figures (from left to right): (1) a visualisation of the significant NBS subnetwork, with the nodes coloured by network and weighted by degree; (2) a heatmap of the proportion of edges within the NBS component that fall within each of the canonical networks as represented quantified using raw (upper triangle) and normalized (lower triangle) proportions (see Supplement for details); and (3) surface renderings depicting the number of edges in the NBS subnetwork attached to each brain region. Vis, Visual network; SomMot, Somatomotor network; DorsAttn, Dorsal Attention network; VentAttn, Ventral Attention network; Lim, Limbic network; FPN, Frontoparietal network; DMN, Default mode network; MTL, medial temporal lobe (amygdala and hippocampus); Thal, Thalamus; Stri, Striatum. Subcortex: PUT, putamen; NAc, nucleus accumbens; CAU, caudate nucleus; GP, globus pallidus; HIP, hippocampus; AMY, amygdala; THA, thalamus; a, anterior; p, posterior; v, ventral; d, dorsal; m, medial; l, lateral; s, superior; i, inferior.

Using raw proportions, connections associated with reduced FC in patients were predominantly concentrated in the default-mode network (Fig 1a). Using normalized proportions, which emphasize network involvement after accounting for differences in network size (see Supplement), limbic and medial-temporal lobe areas were also strongly implicated. At a regional level, the left superior temporal pole, inferior frontal and parietal cortices, and the right post-central gyrus, ventroanterior thalamus, and posterior caudate were among the areas most strongly implicated in the network of FC reductions (Fig 1a). Edges in which FC was higher in patients showed a more homogenous distribution across different networks (Fig 1b). Strongly implicated regions included the right posterior caudate, dorsoanterior and ventroanterior thalamus, left frontal eye field, left cuneus cortex, right anterior caudate, and superior lateral occipital cortex.

#### Baseline to 3-months

We identified a single NBS component showing an altered FC trajectory in the PIPT group compared to controls, comprising 4128 edges linking all 316 regions (*p*_*FWE*_ = .0073). Thus, longitudinal FC changes in antipsychotic-naïve patients were widespread. Within this network, 46% (1913) of edges showed decreasing FC over time and 54% (2215) of edges showed increasing FC in patients over time (Fig 1c-d). The edges showing declining FC over time in PIPT patients had a diffuse distribution across different systems, with some evidence for preferential involvement of FC between the default mode and somatomotor systems (Fig 1c). The right posterior and anterior middle temporal gyrus, dorsoanterior thalamus, bilateral precuneus and posterior cingulate cortex, and left precentral gyrus were among the areas linked to many of the FC reductions over time (Fig 1c).

Edges showing increased FC in PIPT patients over time were strongly concentrated in the DMN, using both raw and normalized counts. Normalized counts additionally implicate the limbic system. Notably, both of these networks showed reduced FC in patients compared to controls at baseline. Regions showing many FC increases over time include right polar and lateral occipital cortices, bilateral post-central, pre-central, and lingual gyri, and left lateral occipital and parietal cortices (Fig 1d).

#### Associations with clinical outcomes

A CCA including 12 principal components each explaining >2% of variance in ΔFC identified a single significant canonical variate linking FC changes to the primary trial outcome measures (Fig 2a; *p*_*FWE*_ = .005, *R* = .901). The canonical loadings indicate that this variate was strongly associated with reducing SOFAS over time (*r* = −.99) and moderately associated with increasing BPRS (*r* = .49). A total of 179 edges showed significant canonical correlations with the ΔFC variate (*p*_*fdr*_ < .05), such that 59 edges correlate positively (fig 2b;. 32 < *r* < .66) and 120 edges correlate negatively (fig 2c; −.41 < *r* < −.65). Positively correlated edges (Fig 2b) largely implicated links between sensorimotor/visual and association and subcortical systems, particularly the striatum (after count normalization). Regions with the largest number of positively correlated edges were the right nucleus accumbens core, left posterior cingulate, and posterior caudate. Negatively correlated edges (Fig 2c) strongly implicated links between somatomotor and visual networks. Regions with the largest number of negatively correlated edges included bilateral pre/post-central gyrus, occipital pole, and lingual gyrus. Thus, across all patients, worse clinical outcome was associated with increased FC between striatal and somatomotor regions, coupled with decreased FC between sensory and motor systems.

**Figure 2.**
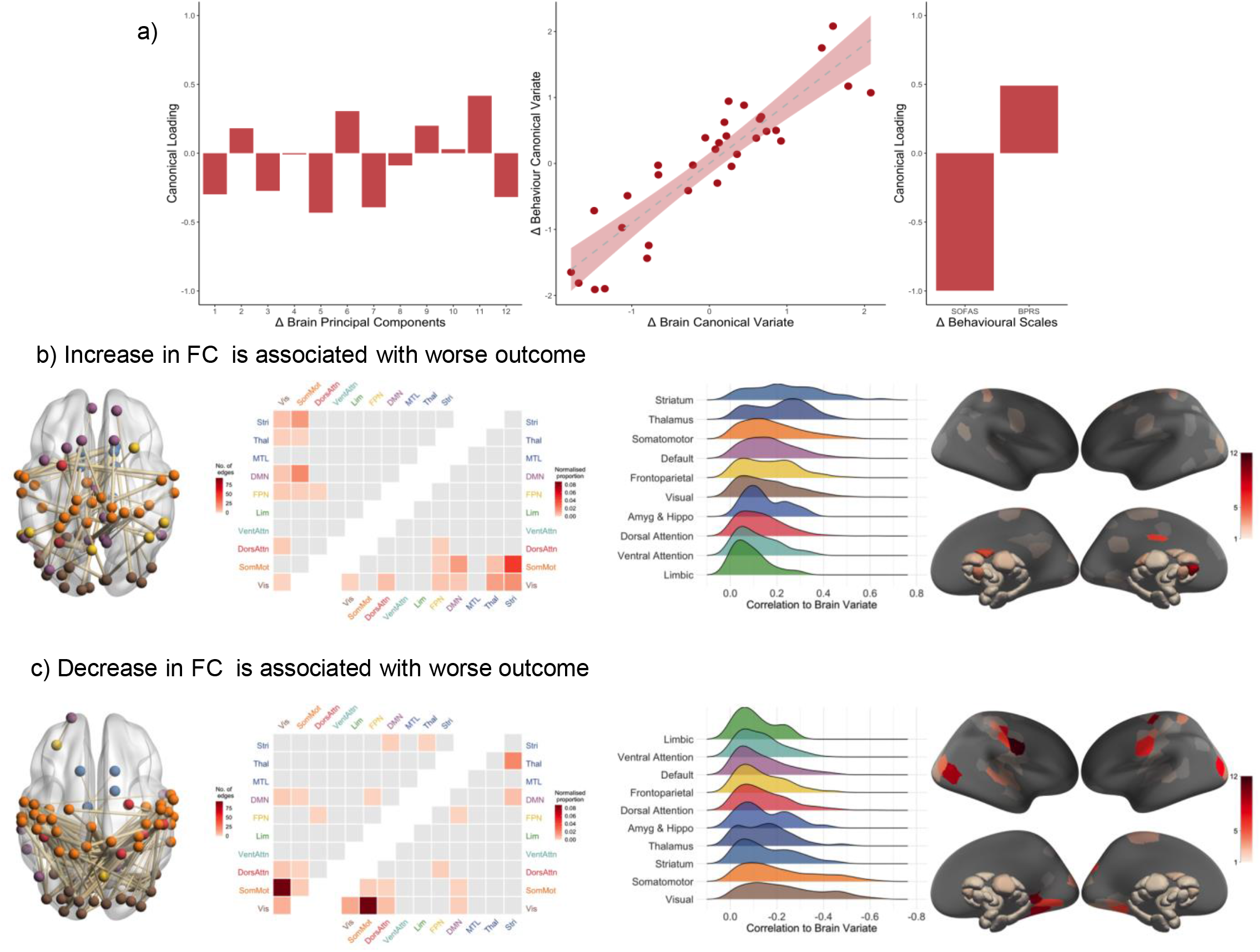
Association between short-term functional connectivity changes and behavioural outcomes in antipsychotic-naïve patients. (A) Canonical loadings of the principal components summarising change in FC (left), scatterplot of the correlation between the brain change and behaviour change canonical variates (middle), and canonical loadings for the change in the two primary trial outcome measures (right; SOFAS, social and occupational functioning assessment scale; BPRS, brief psychiatric rating scale). (B) & (C) depict the edges where change in FC was significantly correlated (FDR-corrected) with the brain change canonical variate. Each of the two panels contains four figures (from left to right): (1) a visualisation of the edges within the NBS component that correlated with the brain change variate, with the nodes coloured by canonical network and weighted by the number of connections to which they are attached in the NBS subnetwork; (2) a heatmap of raw (upper triangle) and normalized (lower triangle) proportions of edges in NBS component that fall within each canonical network; (3) a ridge plot of the distribution of correlations between each edge of the NBS component and the brain variate in the CCA, separated by network and sorted by the median value; (4) surface renderings coloured by the number of edges in the NBS subnetwork attached to each node. Vis, Visual network; SomMot, Somatomotor network; DorsAttn, Dorsal Attention network; VentAttn, Ventral Attention network; Lim, Limbic network; FPN, Frontoparietal network; DMN, Default mode network; MTL, medial temporal lobe (amygdala and hippocampus); Thal, Thalamus; Stri, Striatum.

### Antipsychotic-related changes

#### Baseline to 3-months

We identified a single NBS component showing an altered FC trajectory in the MIPT group compared to the PIPT and control groups, comprising of 634 edges and including all 316 regions (Fig 2; *p*_*FWE*_ = .0001). This network indicates that antipsychotic exposure is generally associated with more FC increases (446 edges) than decreases (188 edges) over time. The subnetwork showing increased FC over time strongly implicates the thalamus and its connections to all other networks, particularly when considering normalized counts (Fig 2a). Regionally, the right lingual gyrus, bilateral occipital pole, left superior frontal gyrus, and right precuneus cortex feature prominently in this subnetwork. Edges showing medication-related FC decreases are more diffusely spread across the networks (Fig 2b). Regionally, decreases predominantly involve the left posterior hippocampus and left inferior frontal gyrus.

## Antipsychotic-related effects

### Association with symptoms and functioning

We find no significant associations between medication-related Δ_3_FC and change in primary outcome measures at 3-months.

#### Longer term Illness- and antipsychotic-related changes (12-months follow-up)

Long-term changes in FC in antipsychotic-naïve patients are circumscribed to a relatively small subset of edges showing large effects concentrated predominantly in the default mode and somatomotor networks (23 edges reduced FC, 10 edges increased FC, linking 30 regions; Sup Fig 2).

Long-term medication-related changes parallel the 3-month results (100 edges reduced FC, 302 edges increased FC, linking 244 regions), being associated with more FC increases than decreases over time, and indicating a preferential involvement of the DMN and limbic systems. Notably, these networks show lower FC in patients at baseline (see Fig 1a).

## Discussion

Our triple-blind, placebo-controlled randomised design allowed us to disentangle longitudinal FC changes in antipsychotic-treated and antipsychotic-naïve patients during the early stages of psychosis. We report evidence for widespread FC changes in antipsychotic-naïve patients within the first few months of an initial psychotic episode, involving both abnormally increasing and decreasing FC over time when compared to healthy controls, with evidence for a preferential involvement and recovery of default mode and limbic systems in patients receiving psychosocial treatment alone. In contrast, antipsychotic-related changes predominantly involve increased FC between the thalamus and other brain regions. Across all patients, longitudinal change in FC over the first three months between subcortical, sensorimotor, and association networks is most strongly related to change in clinical outcome. We additionally report evidence for more circumscribed changes in antipsychotic-naïve patients affecting medial parietal cortex over a 12-month period, with longer-term antipsychotic effects being primarily associated with increased FC in limbic networks.

### Baseline connectivity differences between antipsychotic-naïve patients and control

Our analysis of baseline data identifies a broadly distributed subnetwork of altered FC in antipsychotic-naïve FEP patients, involving an approximately equal proportion of edges showing hyperconnectivity and hypoconnectivity relative to controls. The broad distribution of FC differences we identify is consistent with a general dysconnection hypothesis of psychotic illness^2,36,37^, in which symptoms are tied to a breakdown of inter-regional communication. Our results suggest that the nature of this breakdown, at the outset of illness, is both complex and widespread.

Within this affected subnetwork, FC reductions are predominantly concentrated within the DMN and its interactions with other systems. We also find evidence for preferential involvement of connectivity between the medial temporal lobe (including both the amygdala and hippocampus) and limbic systems with the rest of the brain, after accounting for differences in network size. Together, these findings point to a prominent role for dysconnectivity of limbic and paralimbic systems in FEP. Given the presumed role of the DMN in self-referential and reflective processes^38,39^ and the limbic and medial temporal systems in emotional function^40^, our findings align with views that psychotic symptoms may arise from aberrant salience attributed to internal representations ^41^, as well as evidence for prominent impairments of emotional processing and expression in people with psychotic disorders^42,43^. However, DMN dysfunction is a common finding in many different psychiatric disorders, and may potentially reflect a general vulnerability to psychopathology^44^.

### Connectivity changes in antipsychotic-naïve patients

From baseline to 3 months, we also observe widespread changes in FC in PIPT patients relative to controls. FC reductions are spread relatively evenly across networks, whereas FC increases are strongly concentrated on connections linking the DMN and limbic system to the rest of the brain. Critically, some of these changes in FC, particularly those affecting links between sensory, subcortical, and association networks, are related to improved symptom ratings and functional outcomes over time. It is notable that increased FC is observed in the DMN and limbic systems, which featured prominently in the FC reductions identified at baseline. This result suggests that the dysconnectivity of DMN and paralimbic systems at study intake appears to normalize over the first few months of illness. One explanation for this effect is that patients in the PIPT group received an intensive psychosocial intervention^24,47^. While there is some evidence that such interventions may normalise FC across the limbic system and association networks^48^, previous research has not been able to rule out the effect of antipsychotic medication. Our findings suggest that psychosocial intervention is sufficient to normalise aberrant FC in these systems. Indeed, our analysis of the 12-month time point found far fewer and more circumscribed FC changes over time in the PIPT relative to the control group, suggesting that the majority of FC changes occurred within the first three months of the study, when patients were most intensively engaged with the psychosocial therapy. However, definitively testing this hypothesis requires comparison with an additional treatment group receiving either basic support or no intervention. Such a design is difficult to justify ethically in unmedicated patients.

FC within medial temporal regions was lower at baseline and did not show evidence for preferential increases over time, suggesting that reduced medial temporal FC may represent a core illness-related feature of psychotic illness that is not modified by psychosocial treatment engagement. This result aligns with animal models suggesting a primary role for medial temporal areas in driving the onset of psychotic illness^49^.

### Connectivity changes in antipsychotic-treated patients

Several studies have found evidence of that lowered connectivity at baseline is either normalised or partly normalised after antipsychotic treatment^11,18^ (although see also ^50,51^). Accordingly, we show that antipsychotic exposure largely increases FC and preferentially impacts connections between the thalamus the rest of the brain. The thalamus is a globally connected hub that relays multimodal information between diverse functional networks^52^. The therapeutic efficacy of antipsychotics is primarily mediated by their antagonism of D2-receptors in the striatum^53^, and dopamine dysregulation in psychosis is thought to disrupt striato-thalamo filtering of sensory and limbic information to the cortex^154^. Our findings suggest that antipsychotics may partially remediate communication between thalamus, striatum, and cortex, thereby normalizing information flow within widespread cortico-subcortical systems.

At the 12-month follow-up, we report evidence for a prolonged increase of FC, mainly in the DMN and limbic systems, in medicated patients. This result suggests that an early effect of antipsychotics on thalamo-cortical signalling is followed by a more sustained influence on FC of the DMN and limbic networks, which showed prominent illness-related differences at baseline and a partial recovery, correlated with symptom outcome, in the PIPT group. Normalizing communication between the limbic/paralimbic systems and the rest of the brain may therefore be a primary therapeutic target in FEP.

### Strengths and limitations

The strengths of this study include its prospective randomised control trial design, antipsychotic naïve patients, triple blinding to treatment, and the inclusion of a healthy control group as a reference for characterizing normative change over time. We implemented rigorous and stringent quality control procedures for our rs-fMRI data and used robust non-parametric inference to model longitudinal changes in FC. Our connectome-wide analysis also enables comprehensive mapping of FC differences.

For this study to satisfy ethical concerns, our patients were required to meet strict inclusion criteria related to safety, which may preferentially sample patients with less illness severity. Although the functional and symptom ratings of our patients are comparable to epidemiologically representative or ‘markedly ill’ FEP cohorts^55 56^, we cannot rule out the possibility that patients who remained in the study had a less severe or progressive form of illness than those who did not complete.

## Conclusion

Antipsychotic-naïve FEP patients show widespread functional dysconnectivity at baseline, particularly in limbic and paralimbic systems, with evidence for an improvement of these changes during the first three months of illness. Our results also suggest that antipsychotics may normalise dysconnectivity primarily by impacting in thalamo-cortical and limbic networks.

## Data Availability

Data will be provided upon request and review by the data governance board.

## Supplementary Materials

### Study Design and Funding – Additional Details

The trial took place at the Early Psychosis Prevention and Intervention Centre, which is part of Orygen Youth Health, Melbourne, Australia. The trial was registered with the Australian New Zealand Clinical Trials Registry in November 2007 (ACTRN12607000608460) and received ethics approval from the Melbourne Health Human Research and Ethics committee.

Role of Funding Sources: Janssen-Cilag partially supported the early years of this study with an unrestricted investigator-initiated grant and provided risperidone, paliperidone and matched placebo for the first 30 participants. The study was then funded by an Australian National Health and Medical Research Project grant # 95757. The funders had no role in study design, data collection, data analysis, data interpretation, or writing of this report.

#### Trial Safety Procedures

To further ensure safety, several discontinuation criteria were applied in the clinical trial. These were operationally defined as any of the following: increased risk to self or others (score of ≥5 on the BPRS-4 Suicidality or Hostility subscales, maintained for 1 week); increase in positive psychotic symptom severity (2-point increase on the BPRS-4 subscale of Conceptual Disorganisation, Hallucinations, Unusual Thought Content, or Suspiciousness) maintained for at least 1 week not due to substance use; decrease in overall functioning (20-point drop in SOFAS score from baseline maintained for 1 month); request by the participant for the introduction of antipsychotic medication; failure to satisfactorily recover 3 months after study entry (a score of 5 or more on the BPRS-4 Hallucinations, Suspiciousness, and Unusual Thought Content subscales or a score of 4 or greater on Conceptual Disorganisation); or becoming pregnant. All participants gave written informed consent after having the study fully explained to them, parental consent was also obtained for participants under the age of 18.

#### Antipsychotic and Concomitant Medication Details

Concomitant medications were permitted during the trial, except for additional antipsychotics or mood stabilisers. Four patients within the PIPT group were switched to open-label antipsychotic medication before the 3-month MRI scan and were excluded from the primary analysis. After termination of the randomization phase at 6 months, five patients in the PIPT group were exposed to antipsychotic medication between the 3-month and 12-month scan. Mean cumulative dose and rates of exposure for both patient groups at each timepoint are provided in Supplementary Table 1.

**Supplementary Table 1.** (previously reported^23^) – Cumulative antipsychotic exposure (in Olanzapine milligram equivalates)

|  | Baseline | 3-months | 12-months |
| --- | --- | --- | --- |
| PIPT, mg (M, SD) | 0.16 (0.59) | 78.5 (216)<sup>1</sup> | 608 (1111) |
| MIPT, mg (M, SD) | 1.03 (5.57) | 420 (248) | 1311 (1011) |

<sup>1</sup> Note: In our primary analysis, the 4 patients within the placebo (PIPT) group who were exposed to antipsychotic medication at amounts greater than the study inclusion criteria at the 3-month timepoint were excluded from the primary analysis, thus cumulative antipsychotic exposure of the analysis sample was 4.57mg (14.5)

**Supplementary Figure 1.**
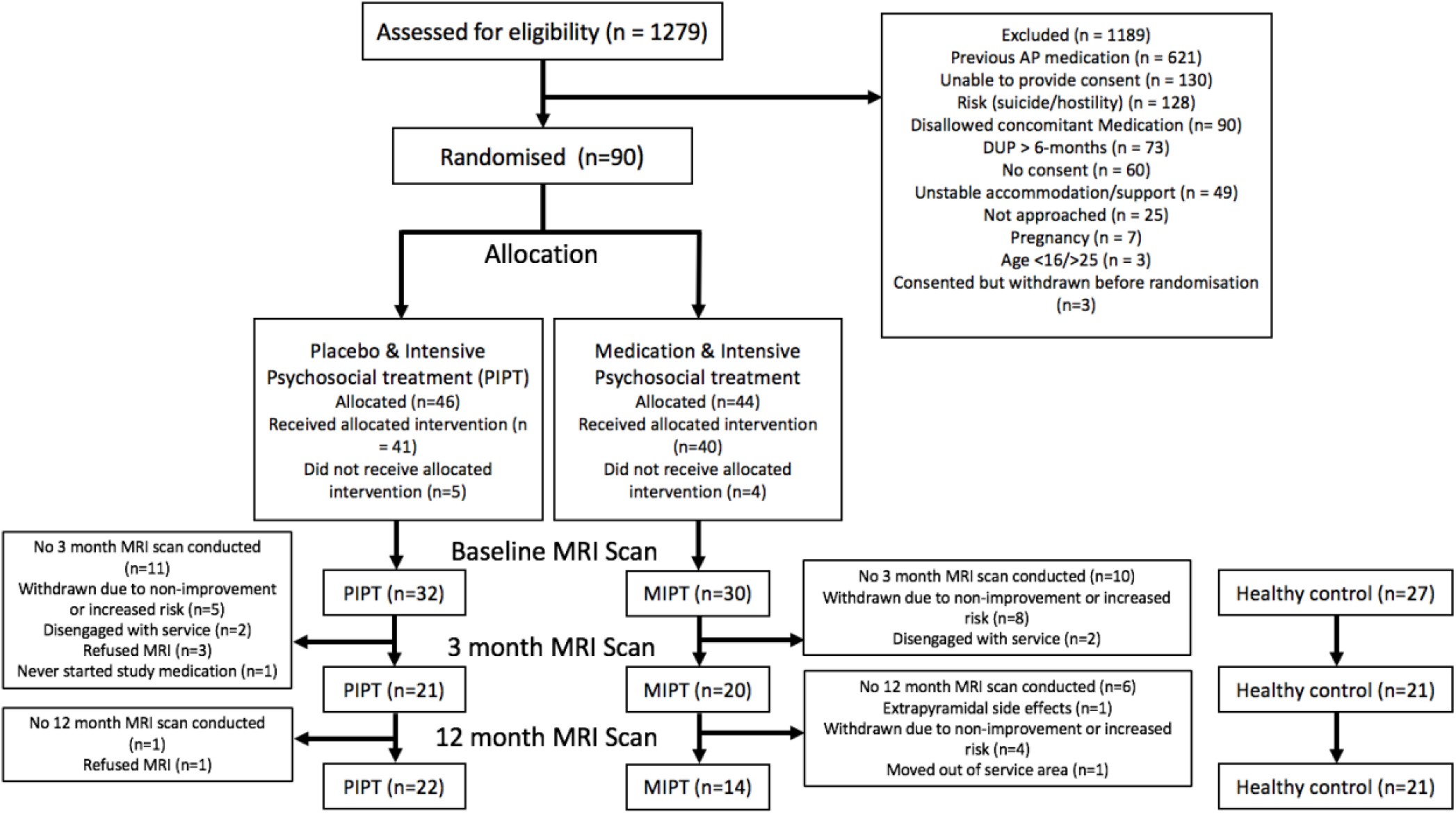
Participant flow diagram.

#### MRI Acquisition Details

Participants were instructed to lie still in the scanner while keeping they eyes closed and maintaining wakefulness. A total of 234 functional volumes with 37 slices each and an interleaved acquisition, were acquired using the following parameters: repetition time = 2000ms; echo time = 32ms; flip angle = 90°; field of view = 210mm; slice thickness of 3.5 mm, and 3.3 x 3.3 x 3.55 mm voxels. For each participant’s T1-weighted image, a total of 176 slices were acquired with an interleaved acquisition using the following parameters: TR = 2300ms; TE = 2.98ms; flip angle of 9°; FOV of 256mm; voxel size of 1.1 x 1.1. x 1.2 mm.

#### Image Processing and Quality Control

A total of 202 rs-fMRI datasets were acquired in this study. Raw images were first put through an automated quality control procedure (MRI-QC)^25^, which resulted in the exclusion of 4 scans due to large artefacts. All remaining images were then processed using a standardised pipeline (*fmriprep;* see Supplement for details)^26^. Briefly, the pipeline included slice time correction, non-linear spatial normalisation to MNI space, brain tissue segmentation, susceptibility distortion estimation and resampling to 2mm^3^. We used previously suggested stringent criteria for excluding and an additional 5 datasets on the basis of excess motion^27^. Automated ICA-based artefact removal (ICA-AROMA)^28^ was applied, then averaged signals from the white matter, CSF, and entire brain were removed from voxelwise time series via linear regression, prior to detrending and high-pass filtering (*f* > .005 Hz). At each stage of pre-processing, quality control (QC-FC) metrics, FC matrices and carpet plots were visualised to ensure the processing step was having the desired effect of mitigating the impact of noise variables. A total of 193 scans survived our quality control procedure. A full quality control report can be accessed here: https://sidchop.github.io/STAGES_rs-fMRI/. To generate whole-brain FC matrices, we parcellated each individual’s normalised scans into 300 cortical^29^ and 32 subcortical regions^30^. We screened all regions for insufficient BOLD signal intensity by first calculating each region’s mean BOLD signal across all 193 scans. We sorted the regional BOLD intensity values from largest to smallest and found the ‘‘elbow’’ of this distribution using the pairwise differences. This led us to exclude 16 cortical parcels, located across the orbitofrontal cortex and temporal pole. In the remainder of the analysis, we therefore used 316 regions. FC was estimated as the Pearson correlation between each pair of regional time series for each individual.

#### Further details on Statistical analysis

Mixed-effects marginal models were used to analyse brain-wide FC changes across the three groups (MIPT, PIPT and controls) and three timepoints (baseline, 3-months and 12-months). At each of the 99,856 edges linking 316 regions, we computed ordinary least squares estimators of group-level regression parameters and a robust-covariance estimator to account for within-subject correlations^31^. This method allows for robust and accurate estimation of random effects while mitigating problems posed by misspecification of covariance structure when using traditional mixed-effects models^31^. The approach also allows for brain-wide non-parametric computation of *p*-values at each edge using wild-bootstrapping^32^ (10,000 bootstraps). All code used to analyse data and generate figures can be accessed here: https://github.com/sidchop/Stages_rs-fMRI/.

The Network Based Statistic (NBS) was used to perform family-wise error-corrected (FWE) inference at the level of connected-components of edges showing a common effect, resulting in a substantial boost in statistical power compared to mass univariate analysis^33^. The NBS procedure involves setting a primary component-forming threshold, *τ* which is applied to both the observed data, and the bootstrap-generated null data. The choice of this threshold is arbitrary; more lenient thresholds will be sensitive to weaker differences distributed over a large number of edges while more stringent thresholds will be sensitive to stronger effects possibly extending over smaller subsets of edges. We report here results for *τ* set to *p* < 0.05 and show results for *p* < 0.01 and *p* < 0.001 in this Supplement. To compute accurate *p*-values for the null data, we used a modified parametric test developed specifically for unbiased inference on marginal models^31^. For both the observed and bootstrap-generated null data, the size (number of edges) of the connected components in the supra-thresholded network was recorded. The size of largest component from each bootstrap was used to build a null distribution and a corrected *p*-value for each observed component was estimated as the proportion of null component sizes that was larger than the observed value.

We evaluated NBS results using a Bonferroni-corrected threshold of *p*_*FWE*_ < 0.016, adjusted for three primary contrasts: (1) a contrast assessing baseline differences between healthy controls and patients; (2) a contrast isolating differential FC changes over time in the antipsychotic-naïve patient (PIPT) group; and (3) a contrast isolating differential FC changes over time in the antipsychotic-treated (MIPT) group. The first contrast maps cross-sectional FC differences in antipsychotic-naïve FEP patients by comparing all patients to controls (neither the MIPT nor PIPT groups had been given antipsychotics at this time). The second contrast was defined as a group by time interaction examining changes in PIPT patients (excluding three patients who were exposed to antipsychotic medication during the treatment period), compared to controls. This maps longitudinal FC changes in FEP patients in the absence of antipsychotic exposure. The third contrast was a conjunction based on the intersection between the group by time interaction for MIPT patients vs controls and the same interaction for MIPT patients vs PIPT patients. This conjunction thus maps FC changes in medicated patients that differ from both antipsychotic-naïve patients and healthy controls to isolate the specific effects of antipsychotic treatment. All contracts were adjusted for age, sex, and mean framewise displacement (head motion).

### Analysis of network level effects

To determine whether the observed FC changes showed any network-specificity, we calculated both the proportion of edges within a given NBS component that fell within each of seven canonical brain networks^29^ (e.g. Fig 1a – upper triangle of matrix). We present both raw proportions and proportions normalized by the total number of possible network connections between each pair of networks (e.g. Fig 1a – lower triangle of matrix); the former identifies preferential involvement of a given network in an absolute sense while the latter accounts for differences in network size (i.e., the tendency for larger networks to be more likely to be implicated in a given NBS network).

We used canonical correlation analysis (CCA) to investigate the relationships between FC changes within any NBS subnetworks and changes in symptoms and functioning across all patients. CCA is a multivariate statistical method that identifies linear combinations of two sets of variables that maximally correlate with each other. The statistical significance of the resulting canonical variates was assessed using permutation-testing^34^. First, we extracted FC estimates for each connection of the sub-network identified in the NBS analysis for both baseline and 3-months timepoints. Residual change scores were computed by regressing 3-months values on baseline values and retaining the residuals (ΔFC). We then summarized these high-dimensional edgewise change values using principal component analysis (PCA), retaining components that account for >2 of variance. Residual change scores for each clinical and functional scale were also computed by regressing 3-months values on baseline values and retaining the residuals. For each of the two identified sub-networks showing longitudinal changes, we computed a CCA assessing the correlation between Δ*FC* and changes in the two pre-registered^24^ primary trial outcome measures: SOFAS and BPRS total scores. The threshold for statistical significance was set to *p*_*FWE*_ < 0.0125, which is Bonferroni-corrected for four CCA analyses which included Δ*F* from each if the primary longitudinal contrasts for both the baseline to 3-months and baseline to 12-months analysis.

To assess which edges contributed most to the ΔFC canonical variate identified in the CCA, we computed the correlation between the residual change score at each edge and theΔFC composite score^35^. To assess statistical significance, we used bootstrapping to estimate the standard error at each edge, computed z-scores by dividing the correlation by the standard error and used these z-scores to compute two-tailed p-values. Edges showing reliable loadings on the canonical variate were those that survived an FDR-correction of these p-values (< .05).

Our secondary analysis repeated the two longitudinal contrasts from our primary analyses, except this time included the 12-month follow-up timepoint, instead of the 3-month timepoint. The contrasts of interest were linear polynomial contrast examining differences in linear trend. We constrain our contrasts in this way, because our hypotheses concern linear interactions between group and time over the follow-up period. The treatment period for the trial ended after 6-months and four PIPT patients commenced antipsychotic medication in this intervening period, in addition to the four patients who had commenced at the 3-month timepoint. Thus, between the 3-month and 12-month scan, a total of eight PIPT patients commenced antipsychotics, whereas all MIPT patients continued medication with varying degrees of exposure. We therefore specified a covariate quantifying cumulative exposure to antipsychotics (olanzapine equivalent, milligrams) for all eight patients within the PIPT group who were exposed to medication at the 3-month or 12-month timepoint. This procedure allowed us to statistically adjust for antipsychotic exposure in the PIPT group when attempting to disentangle the long-term change in FC in antipsychotic-naïve and antipsychotic-treated patients.

## Supplementary Results

### Demographic and sample characteristics

At baseline, the two patient groups (PIPT and MIPT) did not significantly differ in age, education, sex, handedness, BPRS or SOFAS scores. Additionally, the two patient groups did not significantly differ in overall substance use at baseline (*t* = 0.235; *p* = 0. 15) or at 3-months (*t* = −1.36; *p* = 0.1 4). During the trial treatment period, the patient groups did not differ in the rates of antidepressant (χ ^2^ = 0.11; *p* = .732), benzodiazepine (χ ^2^ = 2.94; *p* = .0 6) or other (χ ^2^ = 0.567; *p* = .452) medication exposure.

### Antipsychotic-naïve results (baseline)

Using *τ*-thresholds of *p* < .01 and *p* < .001, we observe that the strongest FC reductions in patients are concentrated in the limbic network and striatum, while the strongest FC increases occur between the thalamus and visual network (Sup. Fig 3).

**Figure 3.**
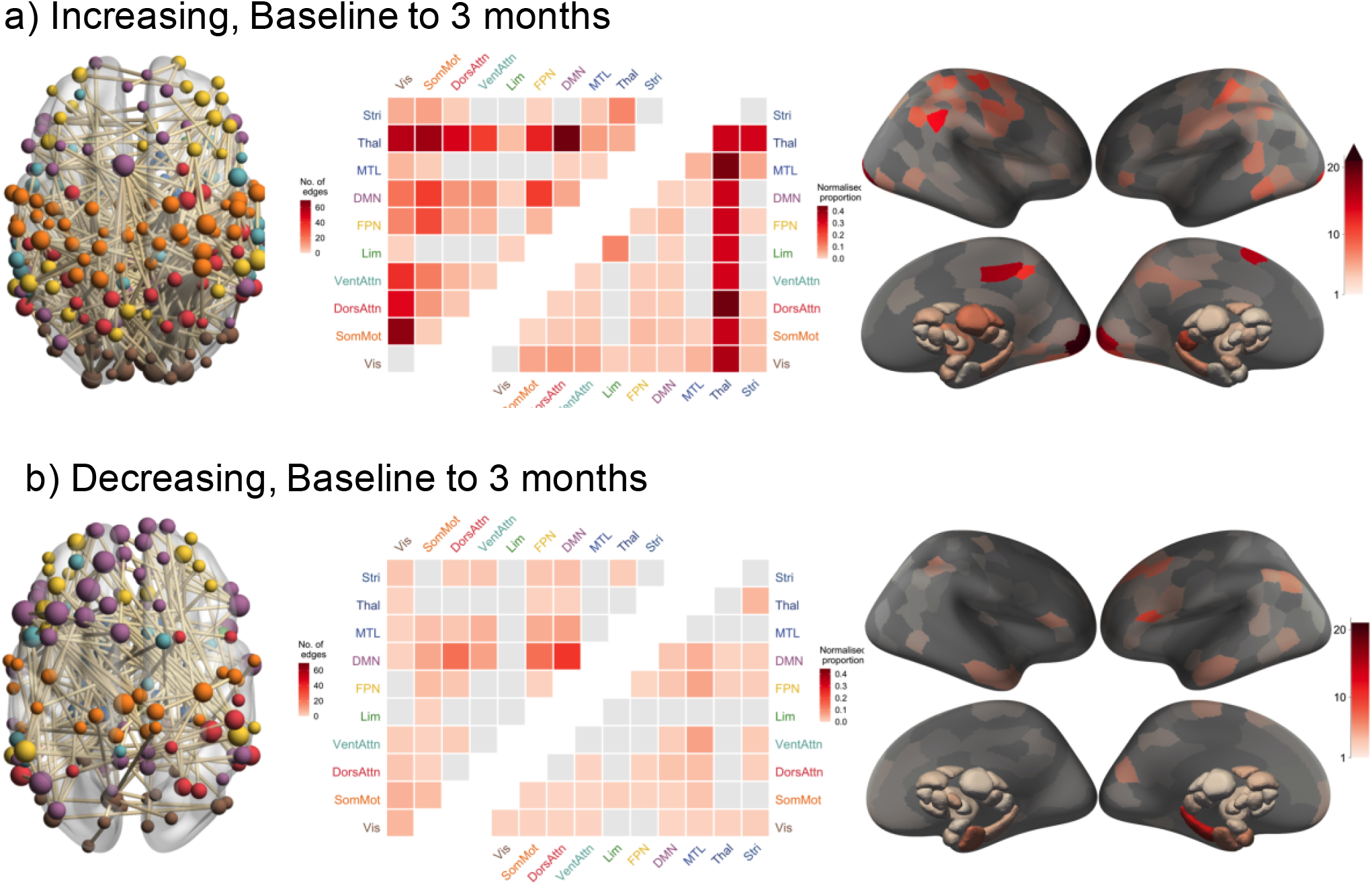
Longitudinal FC changes due to antipsychotic treatment. Each of the two panels contains three figures (from left to right): (1) a visualisation of the significant NBS subnetwork, with the nodes coloured by network and weighted by degree; (2) a heatmap of the proportion of edges within the NBS component that fall within each of the canonical networks as represented quantified using raw (upper triangle) and normalized (lower triangle) proportions (see Supplement for details); and (3) surface renderings depicted the number of edges in the NBS subnetwork attached to each brain region. Vis, Visual network; SomMot, Somatomotor network; DorsAttn, Dorsal Attention network; VentAttn, Ventral Attention network; Lim, Limbic network; FPN, Frontoparietal network; DMN, Default mode network; MTL, medial temporal lobe (amygdala and hippocampus); Thal, Thalamus; Stri, Striatum.

### Antipsychotic-naïve results (baseline to 3-months)

Using *τ*-thresholds of *p* < .01 and *p* < .001, FC decreases in patients remain evenly distributed across networks. Strong FC increases in patients are especially concentrated in the DMN, limbic, and visual networks (Sup. Fig 3).

### Antipsychotic-related results (baseline to 3-months)

Using *τ*-thresholds of *p* < 0.01 and *p* < 0.001, we find evidence that the strongest medication-related FC increases are concentrated in the visual, somatomotor, and attentional networks (Sup. Fig 3). This result aligns with our CCA of antipsychotic-naive changes over time, in which increased ΔFC in sensory networks is associated with improved functioning and symptoms over the first 3 months of illness.

### Long-term Changes in Antipsychotic Naïve Patients compared to healthy controls (Baseline to 12-months)

A significant group-by-time interaction (Sup. Fig 1a-b; p = 0.048_fwe_) was detected only at a *τ*-threshold of p<0.001 (33 edges), with 23 edges showing a decrease and 10 edges showing an increase over 12-months. This result suggests that long-term illness-related changes in FC are circumscribed to a relatively small subset of edges showing large effects. Edges showing reduced FC in patients over time predominantly implicated the default mode and somatomotor networks, with the right precuneus and bilateral post-central being the most heavily implicated brain regions. Edges showing increased FC in patients over time primarily linked DMN, visual, ventral attention and thalamic systems, with the left ventroposterior thalamus and right supra-marginal gyrus being the regions attached to the most edges showing increased FC over time.

### Long-term Antipsychotic-related changes (Baseline to 12-months)

At a *τ*-threshold of p<.05, we identified a single NBS component showing an altered FC trajectory over 12-months in the MIPT group compared to the PIPT and control groups, comprising of 402 edges and including all 316 regions (Sup. Fig 1c-d; p_fwe_ = .0084). As with the 3-month results, medication was associated with a higher number of FC increases (302 edges) over time than decreases (100 edges). Raw counts indicate that these effects were concentrated in limbic, default, frontoparietal, somatomotor and ventral attention networks; normalized counts suggest a preferential concentration in the limbic system. Notably, the limbic network also showed lowered FC in patients at baseline. At a regional level, right prefrontal cortex, and left superior temporal gyrus and posterior hippocampus were strongly implicated in antipsychotic-related increases in FC. Edges showing medication-related decreases in FC were more diffusely spread across the networks, with a strong involvement of the left medial amygdala.

Using *τ*-thresholds of p<0.01 and p<0.001, we find evidence that the strongest effects for medication-related FC increases are concentrated in medial temporal areas and association and attentional networks (Sup Fig 3).

#### Association between Long-term Changes and Symptoms and Functioning

No significant associations were detected between Δ_12_FC in the illness- and medication-related networks and primary outcome measures

**Supplementary Figure 2.**
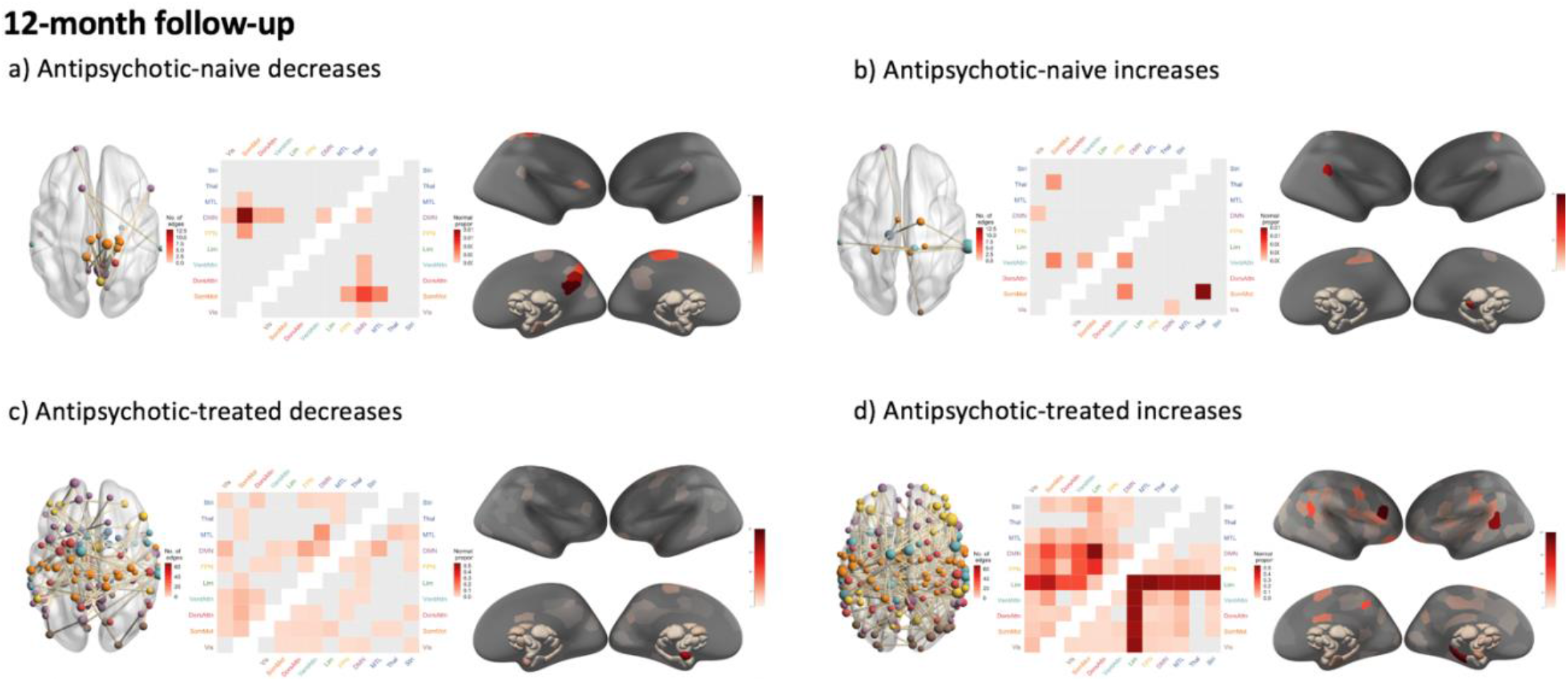
Longer-term effects in antipsychotic-naïve patients (a-b) and longer-term effects related to antipsychotic-medication (c-d) Each of the four panels contains three figures (from left to right): (1) a visualisation of the significant NBS subnetwork, with the nodes coloured by network and weighted by degree; (2) a heatmap of the proportion of edges within the NBS component that fall within each of the canonical networks as represented quantified using raw (upper triangle) and normalized (lower triangle) proportions; and (3) surface renedings depicted the number of edges in the NBS subnetwork attached to each brain region. Vis, Visual network; SomMot, Somatomotor network; DorsAttn, Dorsal Attention network; VentAttn, Ventral Attention network; Lim, Limbic network; FPN, Frontoparietal network; DMN, Default mode network; MTL, medial temporal lobe (amygdala and hippocampus).

**Supplementary Figure 3.**
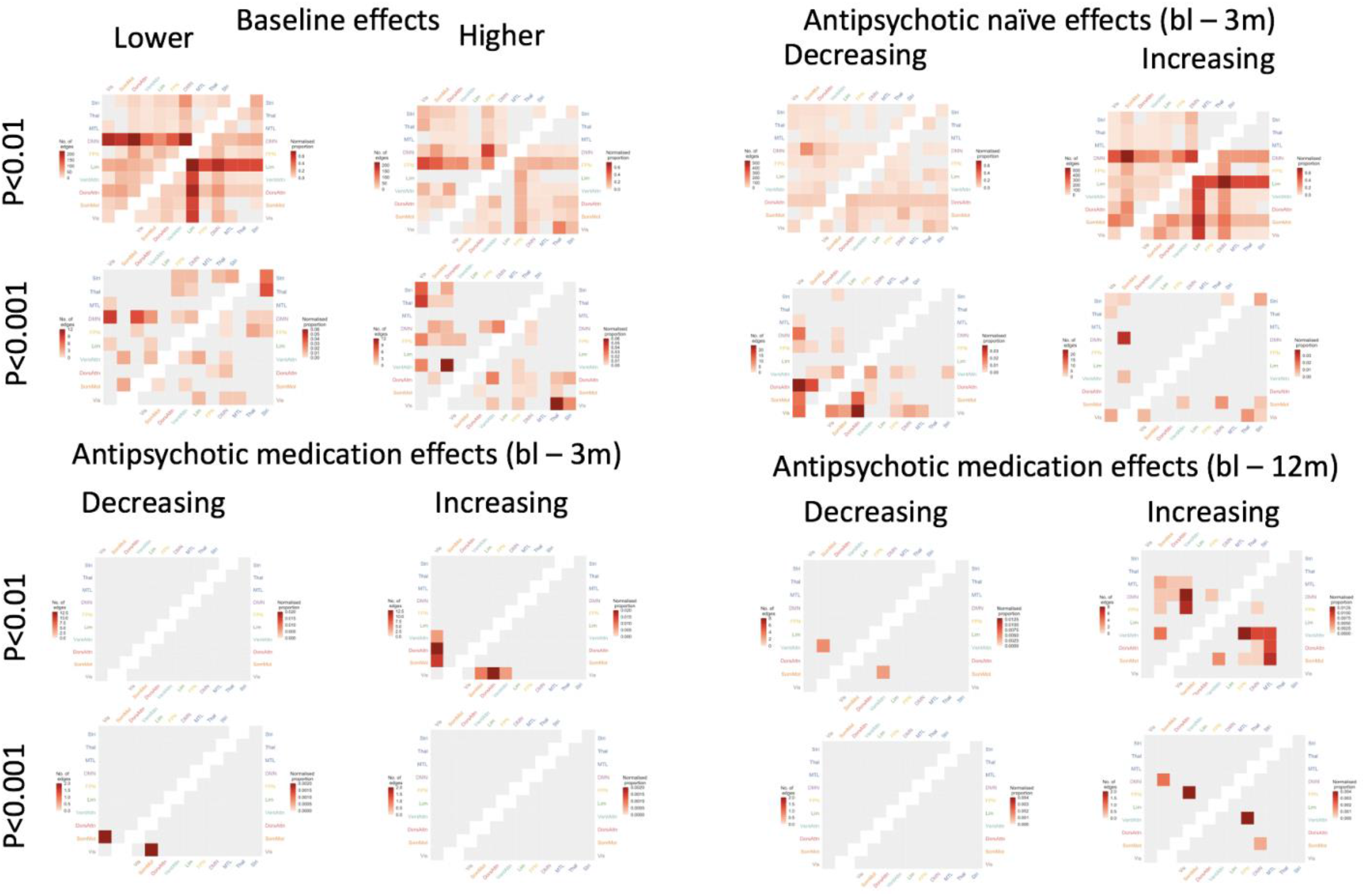
Antipsychotic-naïve and antipsychotic-treated results at a network level (p_fwe_<0.05) at primary thresholds of p <0.01 and p<0.001.

